# “How Autism impacts mothers in a selected autism centre in Karachi, Pakistan- A qualitative study”

**DOI:** 10.64898/2026.09.16.26361740

**Authors:** Aroosa Nighat, Hira Tariq, Fatima Bismah Athar

## Abstract

**Background:** The mothers of children with autism spectrum disorder (ASD) are more stressed than those of neurotypical or any other children with special needs. There remains a notable lack of contextually grounded evidence on the experiences of mothers raising children with autism in resource-constrained settings of a lower middle-income country (LMIC).

**Methodology:** A qualitative phenomenological study was carried out at a government-run autism rehabilitation center over a period of six months. The main purpose of the study was to explore the lived experiences of the mothers of autistic children in a setting like Karachi, Pakistan. Twenty-five mothers of 3-18 years old children enrolled in the selected facility participated in two focus group discussions (FGD) and seven In-depth interviews (IDIs) until data saturation was achieved. Consolidated Criteria for Reporting Qualitative Research (COREQ) guidelines and Guba’s constructs of trustworthiness were followed.

**Results:** Several themes were derived, corresponding to the different layers of the Ecological Systems Theory. Themes included maternal emotional journey, interpersonal relationships, societal perceptions, access to support services and adaptation over time.

**Conclusion:** Mothers of children with Autism in Pakistan face significant challenges at the personal, family, societal, and support systems levels. Removing misconceptions and improving access and quality of services to children with Autism and their families is essential for easing their burden.

**ARTICLE SUMMARY:**

1. This study provides in-depth insight into mothers’ lived experiences in an under-researched LMIC setting.
2. Methodological rigor was enhanced through COREQ, triangulation, member checking, and an audit trail.
3. Conducting the study at a single public-sector center limits transferability. 4-Exclusion of fathers limits broader family perspectives

**What is already known on the topic:** The mothers of children with autism spectrum disorder (ASD) are more stressed than those of neurotypical or any other type of special needs’ children.

Socioeconomic problems, societal stigma and lack of service availability are reported as the major contributors to the caregiver strain experienced by them.

**What this study adds:** The study highlights key needs, barriers, and contextual realities faced by mothers of children with autism within the setting of an LMIC.

**Effect on research, practice or policy:** It can help greatly in managing stigma related to mental health through the perspective of the mothers. Another possible benefit can be the evidence-based formulation of policies and infrastructure required for inclusion of children with ASD into the society.

## INTRODUCTION

Autism Spectrum Disorder (ASD) is a range of life-long neuro-developmental disorders characterized by issues with social communication, restricted interests, and monotonous behaviour. ^(1,2)^ It is the fastest growing neuro-developmental disorder, and its prevalence has dramatically increased up to thirty times since its initial recognition in the late 1960s. ^(3,4)^

The global incidence of autism spectrum disorders according to WHO is 1 in 100 children, while as many as 1 in 35 children in America are reported as being diagnosed with Autism. ^(1,5,6)^ South Asian countries are estimated to have over 5 million of children with ASD. ^(7)^

According to an article published in 2019, India has a reported prevalence of 0.2% which amounts to 2 million autistic people, with half of them in the ‘under 15’ age group. ^(7,8)^ In Pakistan the prevalence of Autism is given as 0.87 %, while unofficial figures give an estimated 350,000 children living with Autism in the country. ^(7,9)^

Approximately 14 million (5.7%) families in the United States are reported to care for a child under 18 years with special needs. ^(10)^ In Europe, the *European Quality of Life Survey* estimates that nearly 100 million individuals, around 20% of the population, are engaged in caregiving across all age groups ^(11).^ In South Asia, the *Carers Worldwide 2024* report estimates approximately 273 million caregivers in India alone. The report further highlights a substantial gender disparity, with 84% of caregivers in India, Nepal, and Bangladesh being women. Additionally, caregiving is associated with considerable personal burden, as 48% of caregivers report concerns about their own health, 89% experience anxiety or depression, and 92% face significant financial challenges. ^(12)^

The mothers of children with autism spectrum disorder (ASD) are known to be more stressed than those of neurotypical or any other type of special needs children. ^(13-15)^ Previous research has highlighted problems in social integration and stigma, family issues, difficulties in the affordability of treatment, lack of professionals having relevant knowledge and deficiency of appropriate services for autism as the major contributors to the caregiver strain experienced by the caregivers of children living with Autism. ^(1,16)^

Research shows that the mother’s religious and socio-cultural background has a strong influence on her capability in dealing with the challenges of raising a child with autism. ^(17)^ Cooperation and support from the families as well as integration in society also play an important part in reducing the stresses associated with looking after a child living with autism spectrum disorder. ^(18,19)^ Parents can also be assisted by providing them with information pertaining to specialized as well as inclusive educational institutions for their autistic child in the vicinity, training them in dealing with their child’s needs in a better way and giving them psychological support to preserve their own mental health. ^(20,21)^

### Rationale

Despite increasing global attention to caregiving, there remains a notable lack of contextually grounded evidence on the experiences of mothers raising children with autism in resource-constrained settings. This gap is particularly pronounced in environments where health, rehabilitation, and social support systems are still evolving. This paper explores the lived experiences of mothers of children with autism, with a focus on understanding the challenges they encounter in caregiving, social integration, and day-to-day navigation of support structures, as well as the coping strategies they employ. By foregrounding maternal perspectives, the study aims to generate nuanced insights that can contribute to strengthening existing support systems and improving responsiveness to their needs. The study is situated within a public-sector autism rehabilitation centre in Karachi, Pakistan, providing an important contextual lens through which these experiences are examined.

### Purpose and Objectives

The study was carried out with the purpose of exploring the lived experiences of mothers of children with autism enrolled in a government run autism rehabilitation centre in Karachi, Pakistan.

The specific objectives were to understand the barriers faced by these mothers in raising and integrating their child into the society and to identify the possible solutions of these problems.

## METHODOLOGY

This study was conducted at the Centre for Autism Rehabilitation and Training Sindh (C-ARTS), a public-sector facility in Karachi, Pakistan. The study population comprised 25 mothers of children aged 3–18 years enrolled at the centre, and data collection was carried out over a period of one month following approval from the Institutional Review Board (IRB).

Consolidated criteria for Reporting Qualitative Research (COREQ guidelines) were followed for the planning, execution, and writing of this qualitative study.^22^The validity and reliability of this study were taken care of by adopting Guba’s constructs of trustworthiness.^23^

The study employed a qualitative phenomenological (empirical) design to explore the lived experiences of mothers of children with autism. A purposive sampling strategy was used to recruit participants with relevant lived experiences. Data was collected through seven in-depth interviews (IDIs) and two focus group discussions (FGDs), with recruitment continuing until thematic saturation was achieved. Mothers of children aged 3 -18 years enrolled at the selected center were eligible to participate, while those who declined to provide informed consent or had a prior history of diagnosed mental illness were excluded.

Data were collected using a semi-structured interview guide designed to elicit detailed narratives on the impact of the child’s condition on the mother’s life, challenges faced in caregiving and social integration, and perceived solutions at multiple levels. The socio-ecological model informed both the development of the interview guide and the analytical framework, enabling exploration of experiences and potential responses at the individual, interpersonal, community, and broader system levels. Participants were physically approached in a naturalistic setting, primarily the waiting areas of the rehabilitation centr. Each in-depth interview (IDI) lasted approximately 20–30 minutes, while focus group discussions (FGDs) ranged from 40–45 minutes. All interviews were audio-recorded with participants’ consent. The Principal Investigator spent approximately one month engaging with participants to build rapport and deepen inquiry. FGDs were conducted in two groups based on the age of the child (<10 years and ≥10 years) to facilitate more context-specific discussions.

A phenomenological approach guided the analysis. Researchers practiced reflexivity by bracketing preconceived assumptions at the outset. All interviews were transcribed verbatim, and data triangulation was achieved through transcripts, field notes, and observational insights. Manual content analysis was conducted using both inductive and deductive approaches, with themes and subthemes mapped onto the domains of the socio-ecological model. Two independent researchers with expertise in qualitative methods conducted the analysis, followed by consensus discussions to ensure rigor and credibility.

The participant public were involved in the conduct and reporting of our research and member checking was performed by sharing the main themes with participants for validation. An audit trail documenting all stages of data collection, coding, and analysis was maintained.

Ethical approval was obtained from the Institutional Scientific Committee and the Institutional Review Board (Approval letter # JSMU/IRB/2023/78), and permission was secured from the administration of the selected institute prior to study initiation. Written informed consent was obtained from all participants. Confidentiality and anonymity were strictly maintained, and the audio recording could only be accessed by the relevant members of the research team. All procedures were conducted in strict accordance with ethical standards, with due respect for participants’ cultural and social contexts.

## RESULTS

Analysis of the qualitative data yielded five overarching themes reflecting the multi-layered experiences of mothers of children with autism. (Fig. 1)

**Figure 1.**
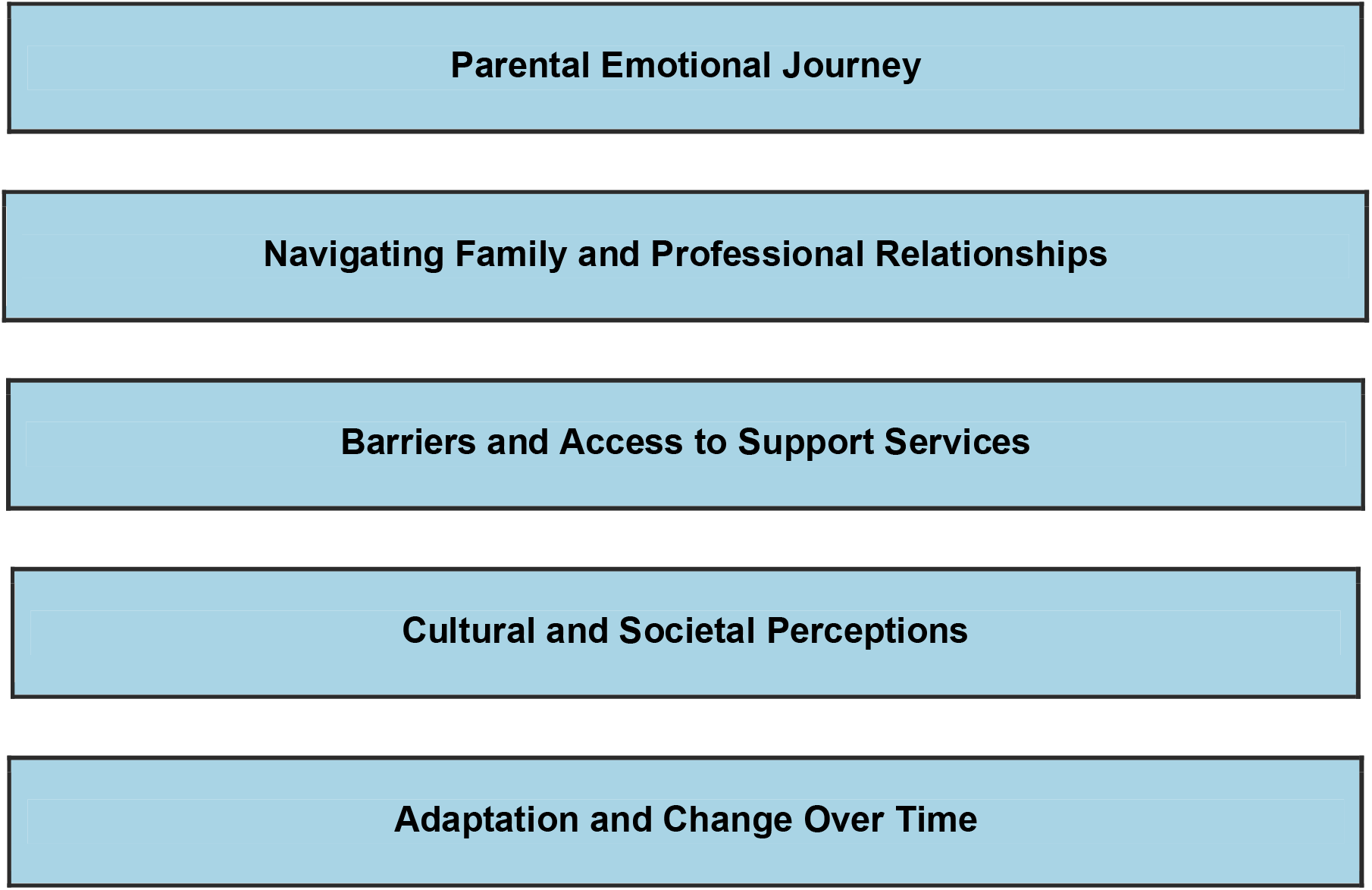
Main themes derived from Bronfenbrenner’s Ecological Systems Theory.

The themes were based on the ‘Ecological systems theory’ by Bronfenbrenner.^24^ This framework provides a holistic view of the multiple contexts influencing a mother’s experiences and challenges. The theory helps to identify the various layers of influence on mothers, ranging from the immediate family dynamics (microsystem) to community support and societal attitudes (macrosystem). The first theme, *parental emotional journey*, captured the evolving psychological responses of mothers at the individual level, ranging from initial distress and uncertainty to gradual acceptance and resilience. The second theme, *navigating family and professional relationships* highlighted the complexities of interpersonal dynamics, including support, conflict, and communication with family members and healthcare providers. The third theme, *barriers and access to support services*, emphasized the systemic and logistical challenges in obtaining timely, affordable, and appropriate care. The fourth theme, c*ultural and societal perceptions* reflected the influence of stigma, social expectations, and community attitudes on maternal experiences. Finally, *adaptation and change over time* illustrated the resilience and coping mechanisms developed by mothers as they adjusted to their caregiving roles corresponding to the ‘chronosystem’ of the ecological systems theory. (Figure 2)

**Figure 2:**
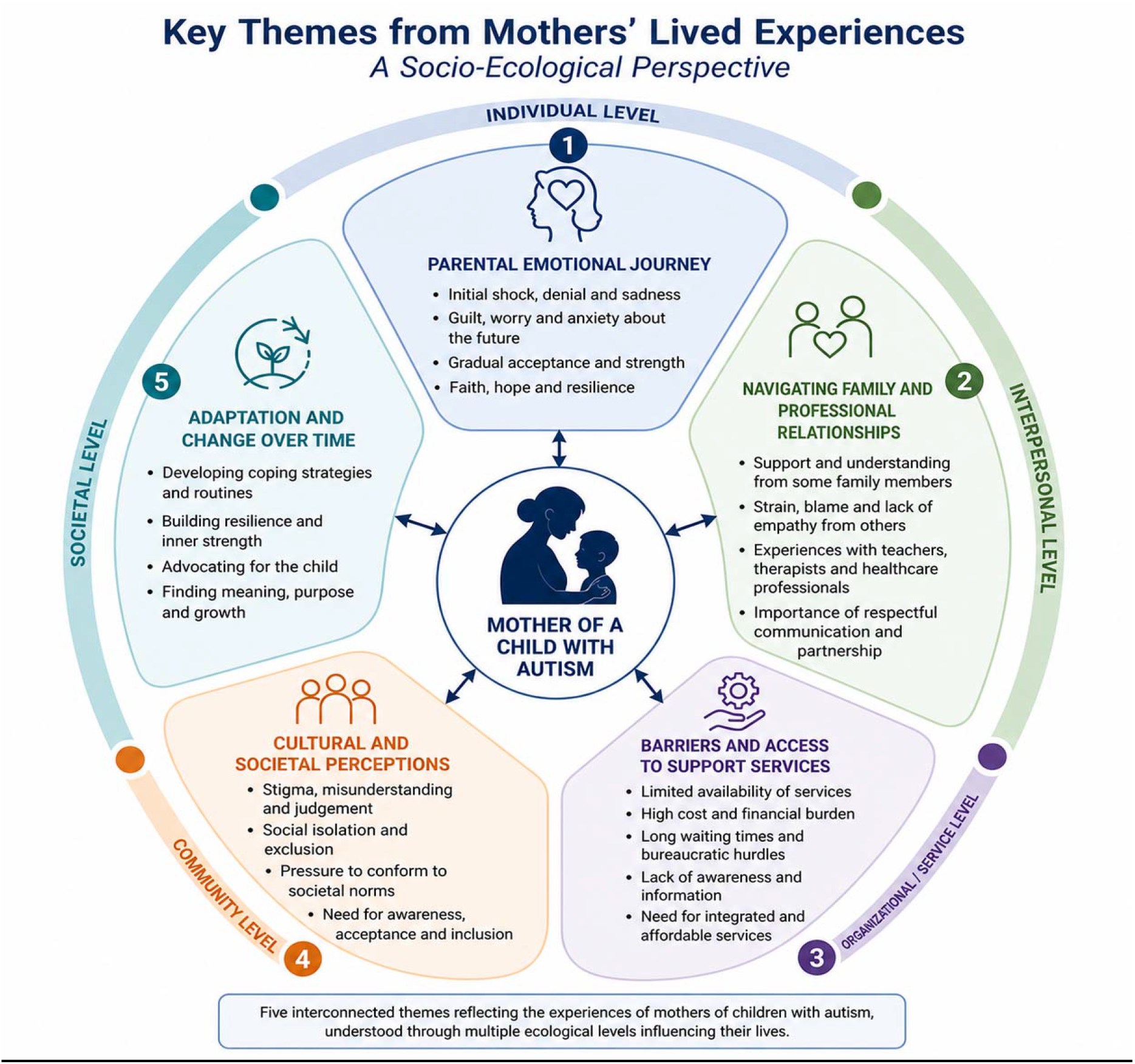
Key themes from mothers’ lived experiences through multiple ecological levels.

### Themes and codes according to the Ecological systems theory

#### Theme 1: Parental Emotional Journey: (Microsystem)

##### Code 1: Personal trauma and Resilience

Mothers expressed a range of emotions from initial shock and confusion to acceptance and advocacy, often shaped by the immediate family environment.

A mother of a child living with Autism expressed her condition on getting the diagnosis as,

> *“I was not mentally stable at all. I was not able to look after my child, neither could I look after myself*.*” (FGD 1, Mother of a 3-10-year-old)*

However, another mother expressed her resilience in these words,

> *“I have spread so much awareness that now people around me are cognizant of autism and know how they can be helpful if they come across another child with Autism*.*” (FGD 2, Mother of an 11-18-year-old)*

##### Code: 2. Daily caregiving challenges

Caring for children with Autism is the biggest challenge that mothers must face every day.

> *“My child even did not have any control on how much he/she was eating neither did he/she have any sense for using the restroom*…*” (IDI 4, Mother of a 3-10-year-old)*

##### Code 3: Personal sacrifices

Most of the mothers were well-educated but had to compromise their professional and family responsibilities while caring for a child with Autism due to the non-availability of any social or systemic support.

A well-qualified mother informed,

> *“I had a job in a reputable organization, but after the diagnosis (of my child), I had to leave it” (IDI 2, Mother of a 3-10-year-old)*

Sometimes, the child with Autism also suffers due to the mother’s other family responsibilities. Mother of another child with ASD shared,

> *“I conceived my second baby when I was attending a child and mother program. I was advised bed rest*… *so I had to leave the program*.*”*

*(IDI 1, Mother of a 3-10-year-old)*

##### Code 4: Process of Diagnosis

Most mothers got concerned regarding the unusual behaviour of their child as early as one month of age, but the formal diagnosis could only be made much later. Usually advise from family or friends or information from the internet prompted parents to seek medical help for their child with autism.

One of the mothers shared her initial observation in these words,

> *“He/She was around a month old when I first doubted that his/her behaviour was a little different from other babies* … *But he/she was properly diagnosed much later*.*” (IDI-2, mother of a 3-10-years-old)*

Another mother of a child living with Autism recalled using internet while searching answers for their child’s unusual behavior,

> *“I searched the internet for his/her symptoms and then came across the term ‘autism’, I matched the symptoms, and all the diagnostic criteria were complete!’ (FGD-1, Mother of a 3-10-year-old)*

#### Theme 2: Navigating Family and Professional Relationships (Mesosystem)

Mothers had to juggle between their responsibilities with the special child and expectations by other family members. Their experiences with the healthcare providers also shaped their response to the situation.

##### Code 1: Family Dynamics and Emotional Support

Family dynamics ranged from conflicts on the diagnosis and management of the child to stigmatizing and blaming the mother for her child’s condition. While positive behaviours included supporting and helping the mother in many ways.

> *“Many people said it’s just because of me, I was the reason of this misfortune*.*” (FGD 2, Mother of an 11-18 -year-old)*

On the other hand, supportive families and friends played a critical role in providing the mother with significant emotional support for the child’s care.

Husbands helped through explaining the child’s condition to the rest of the family, taking care of the children while the mother was busy elsewhere and taking the pressure off her in terms of household chores.

> *“If a father accepts the child, it relaxes the mother, half of her tension are resolved*.*” (FGD 2: mother of an 11-18-year-old)*

The mothers of other children with ASD were also a vital support for these mothers. One of the mothers whose child was enrolled in the Autism centre, explained in these words,

> *“Whenever any mother is feeling depressed, we try to motivate her because only we can understand each other’s problems*.*” (FGD 1, mother of a 3-10-year-old)*

The constant stress associated with looking after a child with Autism often took its toll on the parents’ marital relationship. Mostly the husband blamed the mother for the child’s condition leading to strained relationship between the two. The ordeal was expressed by one of the mothers in these words,

> *“It hurt me so much when (the child with ASD’s) own father resented why (the child with ASD) didn’t die at birth, these things were extremely painful for me*.*” (IDI 7, mother of a 3-10-year-old)*

She also described the challenges frequently faced by the siblings of the child with Autism in these words,

> *“Instead of counselling (child with ASD), I tell my younger child to compromise a little*.*” (IDI 7, mother of a 3-10-year-old)*

##### Code 2: Interaction with healthcare professionals

Seeking professional help also benefited the mother greatly in adjusting to the situation. Mother of a recently diagnosed child explained,

> *“I took sessions from the psychologist (for myself), it greatly improved my mental health and boosted my confidence*.*” (FGD1, Mother of a 3-10-year-old)*

#### Theme 3: Availability and accessibility of support services: (Exosystem)

External factors like the availability of appropriate social services, community resources and policy gaps have a significant impact on the mothers’ experiences and coping mechanisms.

##### Code 1: Experiences with the Healthcare system

In developing countries, the healthcare system is often under-equipped and lacks trained healthcare professionals. One of the mothers shared her experiences as follows,

> *“In Pakistan, there is a lack of specialist doctors there is no proper method of diagnosis It cost us a lot of precious time to get a proper diagnosis for our child*.*” (FGD 2, Mother of an 11-18-year-old)*

In addition, the process of diagnosis and therapies for Autism is very expensive leading to financial difficulties for the family. Another study participant shared her experience in these words,

> *“We had to sell our car for the treatment of our child. As a parent, whatever we can do; we must do it*.*” (IDI 3, Mother of a 3-10-year-old)*

In contrast, a few setups manage these cases professionally resulting in notable improvement in the child’s condition. Another mother appreciated the positive role of healthcare professionals in these words,

> *“When I went to [name of the institute], they started the therapies, and [child’s name] improved a lot. He/she started responding to his/her name and recognizing and naming things*.*” (IDI 6, Mother of a 3-10-year-old child)*

##### Code 2: Experiences with Education/Rehabilitation system

The participant mothers informed us that like the healthcare sector, there was a scarcity of good quality centres for training and rehabilitating children with ASD by the government. Conversely, the private institutes with qualified staff were too costly and difficult to afford. A mother pointed this out in the following way,

> *“As the government-run (rehabilitation) institutions are limited, the private sector is minting money*.*” (FGD 1, Mother of a 3-10-year-old)*

##### Code 3: Policy and advocacy needs

The mothers had a lot to say about the various measures required to improve the lives of kids with Autism. Their suggestions included conducting nationwide surveys to estimate the true burden of autism and other developmental disorders among children. They emphasized having child psychologists in every hospital and mainstream school to facilitate early diagnosis and timely management. They also highlighted the urgent need for more accessible and affordable high-quality centres offering rehabilitation services. For older children they stressed the importance of vocational training and appropriate employment opportunities to make them useful members of society. They also demanded counselling and training facilities for themselves to be better equipped to meet the associated challenges. The study participants further stressed the role of media in reducing the stigma associated with Autism and other mental health disorders through raising public awareness.

One of the mothers made a very pertinent suggestion of finding out the actual burden of disorders among the children at population level, something which is still not known at national level in the country,

> *“A national level survey for any mental health problem with the children can give a complete account of which syndrome, which disorder, or which disease the child has” (IDI 2, mother of a 3-10-year-old)*

She also expressed the need for sufficient government-run centres in these words,

> *“The Government should develop more such institutions, where parents can get the therapies for their children done easily and without excessive financial burden. (FGD 1, mother of a 3-10-year-old)*

Another mother suggested,

> *“Regular trainings should be arranged for the mothers so that they are better equipped to manage any untoward situation at home. (FGD 2, mother of an 11-18-year-old)*

#### Theme 4: Cultural and Societal Perceptions: (Macrosystem)

Broader cultural beliefs and social attitudes influenced mothers’ experiences, making them advocate for better support and awareness regarding Autism.

##### Code 1: Social attitudes and stigma

Children with Autism and their families face significant amount of prejudice and stigma from society. Mothers recognize that the main reason for this is lack of awareness. The mother of a toddler shared,

> “*People think our child is ill-mannered, not adequately raised by the parents. But when told about the child’s condition, they understand that child has a genuine problem” (FGD 1, mother of a 3-10-year-old)*

##### Code 2: Cultural beliefs

The mother of a child enrolled in the centre pointed out,

> *“In our culture, the onus to run the whole household rests entirely on women, on top of it, if she has to oversee a child like the one living with Autism, it further burdens the mother*.*” (FGD 1, mother of a 3-10-year-old)*

#### Theme 5: Adaptation and Change Over Time: (Chronosystem)

Mothers experienced changes in family dynamics and coping strategies over time, influenced by personal growth, increased awareness, and societal changes.

##### Code 1: Evolving family understanding and acceptance

With time, awareness increases, leading to better acceptance in the family. The mother of a child living with ASD explained,

> *Many of the older people in our family don’t understand what autism is. But the younger generation is more accommodating towards it. (FGD 1, mother of a 3-10-year-old)*

##### Code 2: Long-term coping and adaptation strategies

Mothers have used many strategies to deal with this challenge in their lives. These included self-efficacy, self-care, love for the child, sharing the caregiving responsibilities with others, acceptance and ownership, financial planning, seeking better rehabilitation options, religious beliefs and having support groups.

Another mother said,

> *“I now realize that I have to take control of myself; no one can help me in these things*.*” (IDI 2, mother of a 3-10-year-old)*

##### Code 3: Change in the child’s condition over time

During the early years, the families dismiss the possibility of something being wrong with the child due to his early age. But with time, the symptoms become more prominent, convincing the family of unusual behaviour from the child. A mother of a child with Autism described this phenomenon in these words.

> *“People recognize that when an older child is showing odd behaviour, there could be some issue” (FGD 1, mother of a 3-10-year-old)*

However, with an early diagnosis and treatment, mothers acknowledge seeing an improvement in their child. The mother of a child living with ASD explained,

> *“Earlier, he was unable to talk much, but now he can express all his needs*.*’ (FGD 2, mother of an 11-18-year-old)*

## DISCUSSION

This qualitative phenomenological study explored the experiences of mothers of children living with Autism in a resource constrained environment of a lower middle-income country like Pakistan.

The findings were interpreted through the socio-ecological model, which conceptualizes caregiving experiences as shaped by interconnected influences ranging from individual and family dynamics to broader social and structural contexts. Guided by this framework, five key themes emerged: parental emotional journey, navigating family and professional relationships, barriers and access to support services, cultural and societal perceptions, and adaptation and change over time.

Parental emotional journey included elements like personal trauma and resilience in which mothers described their transition from feeling the initial shock and confusion to accepting the situation and even advocating for their child’s rights and spreading awareness among others regarding Autism. An Indian qualitative study published in 2021, also described feeling of shock as an initial reaction in almost all of the respondents, but later adjusting to the situation due to improvement in the child’s condition with professional support.^25^ A systematic review published in 2023 also highlighted the anxiety and poor quality of life due to the challenges faced by the mothers specially the availability and quality of Autism related social support.^26^

Our study also described the role of parental advocacy in shaping their response to their child’s condition. An Australian meta synthesis pointed towards the positive influence of parental advocacy towards better coping experience both for themselves as well as other parents in similar situation. It also emphasized the need for a social support network including primary healthcare workers as well as fellow caregivers in effective management of ASD. ^27^

Parents also talked about the physical, emotional as well as systemic hurdles they must face daily while looking after their child with ASD. Other studies in Singapore and Nigeria also emphasized similar social support related issues faced by the parents of children living with Autism.^28,29^

Our study highlighted the initial sources of information regarding Autism diagnosis as either other people in their social circle or internet, with minimal support from the healthcare system. A recently conducted study in Turkey also reported that the main source of information for these mothers were the electronic and social media and not the scientific and medical resources.^30^

This study highlighted the crucial role of the spouse, immediate family and the parents of other children with ASD in supporting the caregivers. It also reported the beneficial effects of consulting a mental health specialist for coping with this challenge. On the other hand, the mothers also described the adverse effect of the extra demands of caregiving on their relationship with their spouses and other children. Similar findings were reported in a study performed in USA and Greece which concluded that having a child with Autism in the family was negatively associated with healthy marital and family relationships. The study from Greece also highlighted the critical role of family coherence in dealing with this challenge.^31,32^

Mothers also highlighted the challenges in the availability, accessibility and affordability of specialized healthcare services for Autism. A recent systematic review carried out in Iran in 2025 pointed out issues related to timely and adequate diagnosis and lack of information regarding the diagnostic process.^33^ Another systematic review of qualitative studies by Canadian researchers described similar issues with accessibility of diagnostic services for autism worldwide.^34^

Similarly, the mothers complained about lack of specialized educational and training services for the children with ASD. Other studies from India, Iran and South Africa also highlighted similar flaws with their educational system.^25,35,36^

Our research indicated gaps in policy and advocacy for Autism. A case study conducted in Washington DC and published in October 2023 emphasized the benefits of a systematic effort to deal with barriers related to Autism care. The study suggested a multilevel approach involving policy and advocacy for infrastructure development, providing community level support and relevant training to healthcare providers at Primary healthcare level. ^37^

The mothers also described the negative attitudes of society which added to the emotional burden experienced by these mothers. Similar thoughts were expressed in some recent studies conducted in India and Nigeria where the participants described the social rejection and inconsiderate behaviour of the social circle of these mothers.^25, 29^ A systematic review published in 2025 pointed towards strong links between caregiver strain and the social stigma faced by the caregivers of children with ASD. It also highlighted the positive effect of a supportive social circle in dealing with the stigma and its associated psychosocial impact.^38^ Another study from Quebec, Canada reaffirmed these findings and called for policies and procedures focused on preventing the societal prejudice faced by the family members of individuals with ASD. ^39^

The mothers of children living with Autism described the healing effect of time on the acceptance of the child’s condition by themselves as well as the rest of the family. The child’s condition also improved over time following proper diagnosis and management of their disorder. A review article published in 2023 also highlighted the positive outcomes of early diagnosis and appropriate management both on the progress of the child with ASD and acceptance of his condition by his family members. However, the review also warned against the stigma and mental anguish these families must face following such a diagnosis. The study emphasized accurate diagnosis, assistance to the families, and promotion of societal awareness as measures to counteract these issues. It also suggested the use of latest technologies such as biomarkers and artificial intelligence to avoid misdiagnosing Autism in such children.^40^

### Strengths, Limitations and Recommendations

This study provides an in-depth exploration of the lived experiences of mothers of children with autism within a developing country context, offering valuable insights into an under-researched population. The use of the socio-ecological model enabled a systematic understanding of how multiple environmental layers from individual and family dynamics to broader societal influences shape maternal experiences and caregiving challenges. The study further highlights key needs, barriers, and contextual realities faced by mothers, contributing evidence that can support the strengthening of existing support systems and service delivery for children with autism and their families. However, certain limitations should be acknowledged. The study was conducted at a single public-sector rehabilitation centre, which may limit the transferability of findings to other settings. Additionally, the perspectives of mothers whose children were not enrolled in any rehabilitation services were not captured, and the exclusion of fathers’ experiences limits a more comprehensive understanding of family dynamics.

Addressing the challenges identified in this study requires coordinated, multi-level efforts. There is a need for nationally representative data to better estimate the burden of autism and related developmental disorders. Routine screening at primary healthcare facilities and early childhood education centres should be strengthened to enable early diagnosis and timely intervention. Training healthcare professionals at primary care level and using the latest diagnostic techniques such as biomarkers and use of diagnostic AI tools can be instrumental in achieving this. Expanding access to affordable, high-quality rehabilitation services through greater public-sector investment is essential. In addition, structured vocational training and employment opportunities for older children and adolescents with autism should be developed to promote long-term independence and reduce familial burden. Strengthening counselling and support services for parents and families is equally important to enhance coping capacity. Media and community-based initiatives can play a significant role in raising awareness and addressing stigma associated with autism.

### Conclusion

In conclusion, the mothers of children with autism navigate complex, multi-layered challenges that extend beyond the immediate caregiving environment into broader social and structural domains. Their experiences highlight the need for more responsive, inclusive, and accessible support systems. Addressing these gaps through strengthened services, improved awareness, and contextual relevant interventions can contribute to improving the well-being of the children with autism as well as their families.

## Data Availability

All relevant data produced in the present work are contained in the in the manuscript. Raw data produced in the present study are available upon reasonable request to the authors.

## Acknowledgements

The authors would like to acknowledge the administration and staff of ‘Centre of Autism Rehabilitation and Training-Sindh’ for their cooperation in carrying out this research.

## Author Contributions

Dr Aroosa Nighat (AN): Corresponding author. Conceived the idea, developed the proposal, performed data acquisition, analysis and interpretation. Prepared the first and final draft of the manuscript. Responsible and accountable for the accuracy or integrity of the work.

Dr Hira Tariq (HT): Performed analysis and reviewed the manuscript critically for the final draft. Responsible and accountable for the accuracy or integrity of the work.

Ms. Fatima Bismah Athar (FBA): Assisted in data acquisition, performed literature search and drafted some sections of the article, reviewed the manuscript critically for the final version to be published. Responsible and accountable for the accuracy or integrity of the work.

